# Trends in clinical characteristics and associations of severe non-respiratory events related to SARS-CoV-2

**DOI:** 10.1101/2021.03.24.21251900

**Authors:** Tal El-Hay, Ehud Karavani, Asaf Peretz, Matan Ninio, Sivan Ravid, Michal Chorev, Michal Rosen-Zvi, Tal Patalon, Yishai Shimoni, Anil Jain

## Abstract

**Background:** The 2019 novel coronavirus (SARS-CoV-2) is reported to result in both respiratory and non-respiratory severe health outcomes, but quantitative assessment of the risk – while adjusting for underlying risk driven by comorbidities – is not yet established.

**Methods:** A retrospective observational study using electronic health records of 9,344,021 individuals across the U.S. with at-least 1 year of clinical history and followed up throughout 2020.

**Results:** 131,329 individuals were associated with SARS-CoV-2 infection by January 6, 2021 in three distinct surges. While the age and number of preexisting conditions had decreased throughout the pandemic, the characteristics of those who experienced severe health events did not.

During the second surge, between June 7 and November 18, 2020, 425,988 individuals in the base cohort were admitted to emergency rooms or hospitals. Among them, 15,486 were detected with SAR-CoV-2 within few days of admission. Significant adjusted odds ratios were observed between SARS-CoV-2 infection and the following severe health events: respiratory (4.38, 95% confidence interval 4.16– 4.62), bacterial pneumonia (3.25, 2.76–3.83), sepsis (1.71, 1.53–1.91), renal (1.69, 1.57–1.83), hematologic/immune (1.32, 1.20–1.45), neurological (1.23, 1.09–1.38).

**Conclusions:** SARS-CoV-2 infection among hospitalized patients is associated with non-negligible increased risk of severe events including multiple non-respiratory ones. These associations, which complement recent studies, are persistent even after accounting for sources of selection and confounding bias, increasing the confidence they are not spurious.

## Introduction

Coronavirus disease 2019 (COVID-19), caused by the severe acute respiratory syndrome coronavirus 2 (SARS-CoV-2), has emerged in Wuhan, China in late 2019 ^1^. The virus then rapidly spread worldwide, causing the first known case in the United States on January 20^th^, 2020 ^2^, and subsequently infecting millions throughout the year ^3^.

SARS-CoV-2 is a respiratory virus, but its disease is not limited to the respiratory system. While most COVID-19 cases suffer from mild common-cold-like symptoms to severe respiratory disorders, a growing body of evidence suggests the disease can also cause multi-organ dysfunction – with patients manifesting cardiac, hepatic, renal, and neurological conditions. However, access to quantitative, risk-adjusted comparison of incidence of such events with non-infected populations is still scarce.

Time is a major confounding factor in emerging diseases, since care evolves over time and susceptible population may change. Therefore, there is also a need to characterize the pandemic over time. Prior studies either described clinical characteristics in a relatively small sample and a single time point ^4,5,6^, or used large-sample state-bureaus data that lack clinical background. Recent works started using electronic health records that allow both larger sample sizes and a clinical history to diagnose COVID-19 ^7^, to describe adverse events ^8^ and to infer demographic risk factor ^9^.

In this work we use the IBM Explorys database to analyze detailed clinical and demographic data from multiple centers across the United States. Being large in both breadth and depth, the Explorys database provides a diverse sample with a nation-wide coverage of hospitals and other healthcare facilities that is also rich with detailed patients’ characteristics. It contains information on those detected with SARS-CoV-2 and a general baseline population. This longitudinal database is ideal to address our two main goals: a) to describe temporal changes of individuals detected with SARS-CoV-2 and b) to quantify adjusted associations between SARS-CoV-2 infection and severe health events in multiple organ systems.

## Results

### Cohort characteristics at baseline

We define a *base population cohort* to be composed of individuals who had some record in the database for both 2019 and 2020 (Figure 1). Intotal the base population cohort includes 9,344,021, out of which 131,329 have had a SARS-CoV-2 diagnosis before January 6, 2021 and are denoted the *base detected cohort*. The base detected cohort is, on average, 4 years older than the base population cohort and with lower variance (Table 1, Supp-Figure 1). Other demographic factors are similar between the two cohorts, except for the relative over-representation of (self-reported) African Americans and Hispanics among those detected with SARS-CoV-2, which is consistent with previous reports of racial disparities in the US ^10,11^. Clinical factors also exhibit bias, where individuals in the base detected cohort tend to have higher rates of various preexisting conditions such as cardiac disorders, diabetes, and neurological disorders (Table 1). Similarly, those detected with SARS-CoV-2 show higher incidence of severe health events previously reported to be presented by COVID-19 patients ^12^ (see Methods).

**Table 1.** Characteristics of participants in the base population, separated to those detected with SARS-CoV-2 and the rest.

|  | General Population |  | SARS-2 diagnosed |  |
| --- | --- | --- | --- | --- |
|  | n=9,212,692 | % | n=131,329 | % |
| Demographics |  |  |  |  |
| Age (Mean, STD) | 45.0 | 24.4 | 49.3 | 20.6 |
| Missing | 1,154 | 0.0% | 0 | 0.0% |
| Sex |  |  |  |  |
| Female | 5,015,598 | 54.4% | 78,335 | 59.6% |
| Male | 3,726,744 | 40.5% | 52,259 | 39.8% |
| Missing | 470,350 | 5.1% | 735 | 0.6% |
| Race |  |  |  |  |
| African American | 1,159,877 | 12.6% | 30,245 | 23.0% |
| Asian | 131,240 | 1.4% | 1,232 | 0.9% |
| Caucasian | 5,775,200 | 62.7% | 89,499 | 68.1% |
| Hispanic/Latino | 49,442 | 0.5% | 112 | 0.1% |
| Multi-racial | 124,424 | 1.4% | 2,154 | 1.6% |
| Other | 169,013 | 1.8% | 1,695 | 1.3% |
| Refused to classify | 98,309 | 1.1% | 1,360 | 1.0% |
| Missing | 1,705,187 | 18.5% | 5,032 | 3.8% |
| Ethnicity |  |  |  |  |
| Declined | 173,764 | 1.9% | 1,181 | 0.9% |
| Hispanic | 419,872 | 4.6% | 6,129 | 4.7% |
| Non-Hispanic | 5,792,895 | 62.9% | 77,782 | 59.2% |
| Other | 830,950 | 9.0% | 39,001 | 29.7% |
| Missing | 1,995,211 | 21.7% | 7,236 | 5.5% |
| Smoking |  |  |  |  |
| Current | 763,108 | 8.3% | 9,944 | 7.6% |
| Former | 1,020,434 | 11.1% | 24,949 | 19.0% |
| Never | 2,894,037 | 31.4% | 64,589 | 49.2% |
| Missing | 4,535,113 | 49.2% | 31,847 | 24.2% |
| Insurance |  |  |  |  |
| Medicaid | 938,590 | 10.2% | 11,249 | 8.6% |
| Medicare | 1,244,038 | 13.5% | 18,144 | 13.8% |
| Other | 116,646 | 1.3% | 1,342 | 1.0% |
| Other public | 54,657 | 0.6% | 1,078 | 0.8% |
| Private | 3,859,656 | 41.9% | 54,155 | 41.2% |
| Self-pay | 696,710 | 7.6% | 18,267 | 13.9% |
| Missing | 2,999,105 | 32.6% | 45,361 | 34.5% |
| Comorbidities |  |  |  |  |
| Alcohol substance abuse | 323,241 | 3.5% | 5,835 | 4.4% |
| Cardiac disorder |  |  |  |  |
| Acute MI | 90,125 | 1.0% | 2,322 | 1.8% |
| Carditis cardiomyopathy | 139,467 | 1.5% | 3,474 | 2.6% |
| Cardiac arrest | 20,161 | 0.2% | 397 | 0.3% |
| CHF | 290,149 | 3.1% | 8,135 | 6.2% |
| Conduction | 244,863 | 2.7% | 5,007 | 3.8% |
| Coronary | 543,135 | 5.9% | 11,662 | 8.9% |
| Dysrhythmias | 924,163 | 10.0% | 21,870 | 16.7% |
| Pulmonary | 152,410 | 1.7% | 4,549 | 3.5% |
| Valve disorders | 360,635 | 3.9% | 6,771 | 5.2% |
| Cerebrovascular disease | 361,378 | 3.9% | 9,137 | 7.0% |
| Delirium dementia cognitive disorders | 143,872 | 1.6% | 4,972 | 3.8% |
| Diabetes | 1,433,843 | 15.6% | 35,840 | 27.3% |
| Epilepsy and convulsions | 167,414 | 1.8% | 3,314 | 2.5% |
| Gastrointestinal disorders | 928,769 | 10.1% | 22,842 | 17.4% |
| Hematologic disorders | 923,752 | 10.0% | 25,694 | 19.6% |
| Hematologic neoplasms | 89,123 | 1.0% | 2,215 | 1.7% |
| Hepatitis | 98,424 | 1.1% | 2,127 | 1.6% |
| HIV | 19,584 | 0.2% | 411 | 0.3% |
| Hypertension | 2,395,126 | 26.0% | 53,163 | 40.5% |
| Immunity disorders | 36,800 | 0.4% | 1,510 | 1.1% |
| Lipid metabolism disorders | 2,133,132 | 23.2% | 45,975 | 35.0% |
| Liver disorders | 393,844 | 4.3% | 11,228 | 8.5% |
| Malignant solid neoplasms | 853,082 | 9.3% | 17,807 | 13.6% |
| Migraine | 392,025 | 4.3% | 10,941 | 8.3% |
| Multiple sclerosis | 36,445 | 0.4% | 801 | 0.6% |
| Musculoskeletal connective tissue disorders | 242,582 | 2.6% | 6,069 | 4.6% |
| Neurological disorders | 661,537 | 7.2% | 19,058 | 14.5% |
| Nutritional deficiencies | 1,059,362 | 11.5% | 30,454 | 23.2% |
| Obesity | 1,612,466 | 17.5% | 43,760 | 33.3% |
| Pancreatic disorders | 73,740 | 0.8% | 1,831 | 1.4% |
| Parkinson's disease | 35,322 | 0.4% | 790 | 0.6% |
| Peripheral and visceral vascular disease | 339,951 | 3.7% | 8,474 | 6.5% |
| Psychiatric disorders | 1,751,388 | 19.0% | 39,542 | 30.1% |
| Pulmonary disorder |  |  |  |  |
| Asthma | 655,241 | 7.1% | 14,240 | 10.8% |
| COPD bronchiectasis | 172,842 | 1.9% | 4,362 | 3.3% |
| Lung disease due to external agents | 8,658 | 0.1% | 266 | 0.2% |
| Pleurisy pneumothorax | 114,527 | 1.2% | 2,886 | 2.2% |
| Pulmonary fibrosis | 35,391 | 0.4% | 1,026 | 0.8% |
| Respiratory failure | 189,565 | 2.1% | 9,805 | 7.5% |
| Renal disorders | 591,292 | 6.4% | 16,909 | 12.9% |
| Respiratory infections | 600,077 | 6.5% | 17,730 | 13.5% |
| Skin infections | 395,529 | 4.3% | 9,564 | 7.3% |
| Thyroid disorders | 894,547 | 9.7% | 19,499 | 14.8% |
| Outcomes |  |  |  |  |
| Cardiovascular | 76,627 | 0.8% | 4,478 | 3.4% |
| Hematologic immune | 82,361 | 0.9% | 5,496 | 4.2% |
| Hepato | 3,429 | 0.0% | 177 | 0.1% |
| Neurological | 72,621 | 0.8% | 3,913 | 3.0% |
| Renal | 87,813 | 1.0% | 7,841 | 6.0% |
| Respiratory bacterial pneumonia | 9,705 | 0.1% | 1,442 | 1.1% |
| Respiratory | 194,656 | 2.1% | 21,442 | 16.3% |
| Respiratory viral pneumonia | 1,115 | 0.0% | 11,771 | 9.0% |
| Sepsis | 32,595 | 0.4% | 3,799 | 2.9% |
| **Any** | 349,038 | 3.8% | 25,929 | 19.7% |
| **None** | 8,863,654 | 96.2% | 105,400 | 80.3% |
| **Treatments** |  |  |  |  |
| **Anticoagulants** | 243,151 | 2.6% | 12,090 | 9.2% |
| **Convalescent plasma therapy** | 1,493 | 0.0% | 664 | 0.5% |
| **Cytokine inhibitors** | 1,059 | 0.0% | 346 | 0.3% |
| **Dexamethasone (and other corticosteroids)** | 429,778 | 4.7% | 21,935 | 16.7% |
| **Hydroxychloroquine** | 20,454 | 0.2% | 2,303 | 1.8% |
| **Remdesivir** | 203 | 0.0% | 6,656 | 5.1% |
| **Tracheostomy** | 3,038 | 0.0% | 243 | 0.2% |
| **Ventilation moderate** | 53,766 | 0.6% | 4,476 | 3.4% |
| **Ventilation severe** | 7,209 | 0.1% | 1,405 | 1.1% |
| **Ventilators and other respiratory support devices** | 20,715 | 0.2% | 1,945 | 1.5% |

**Figure 1.**
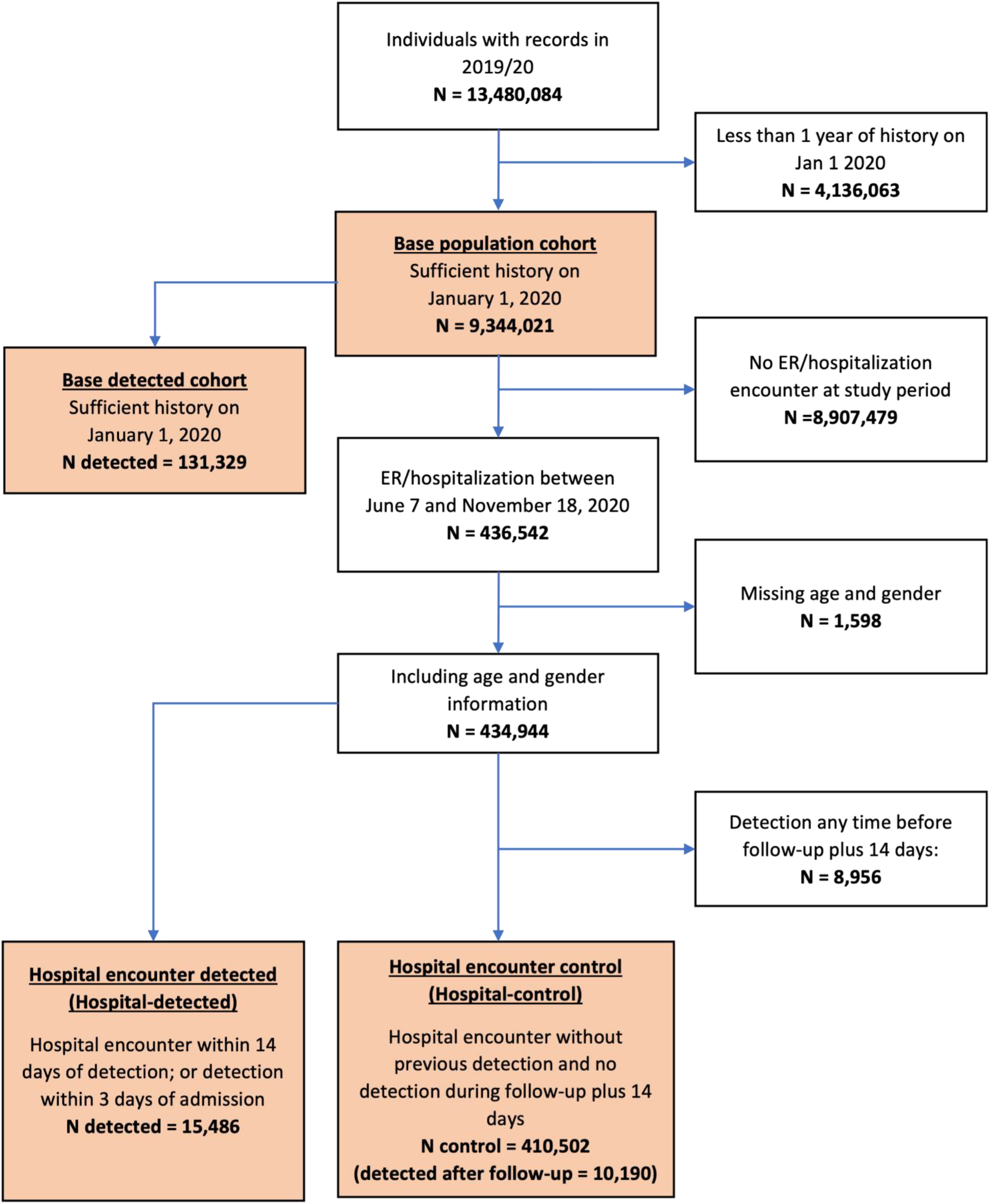
Flow (STROBE) diagram depicting participants in the study. Populations used in the study are colored with orange. The temporal analysis in the study uses the Base population cohort and the association analysis uses the Hospital encounter cohor t.

### Temporal patterns of infection detection

We followed temporal patterns in the base detected cohort over the period of time between March 2020 and December 2020. The lower panel of Figure 2 shows the weekly number of diagnosed individuals, which exhibits three surges of SARS-CoV-2 infection detections: the first starting in mid-March; the second one starting in early June; and a third one starting mid-October.

**Figure 2.**
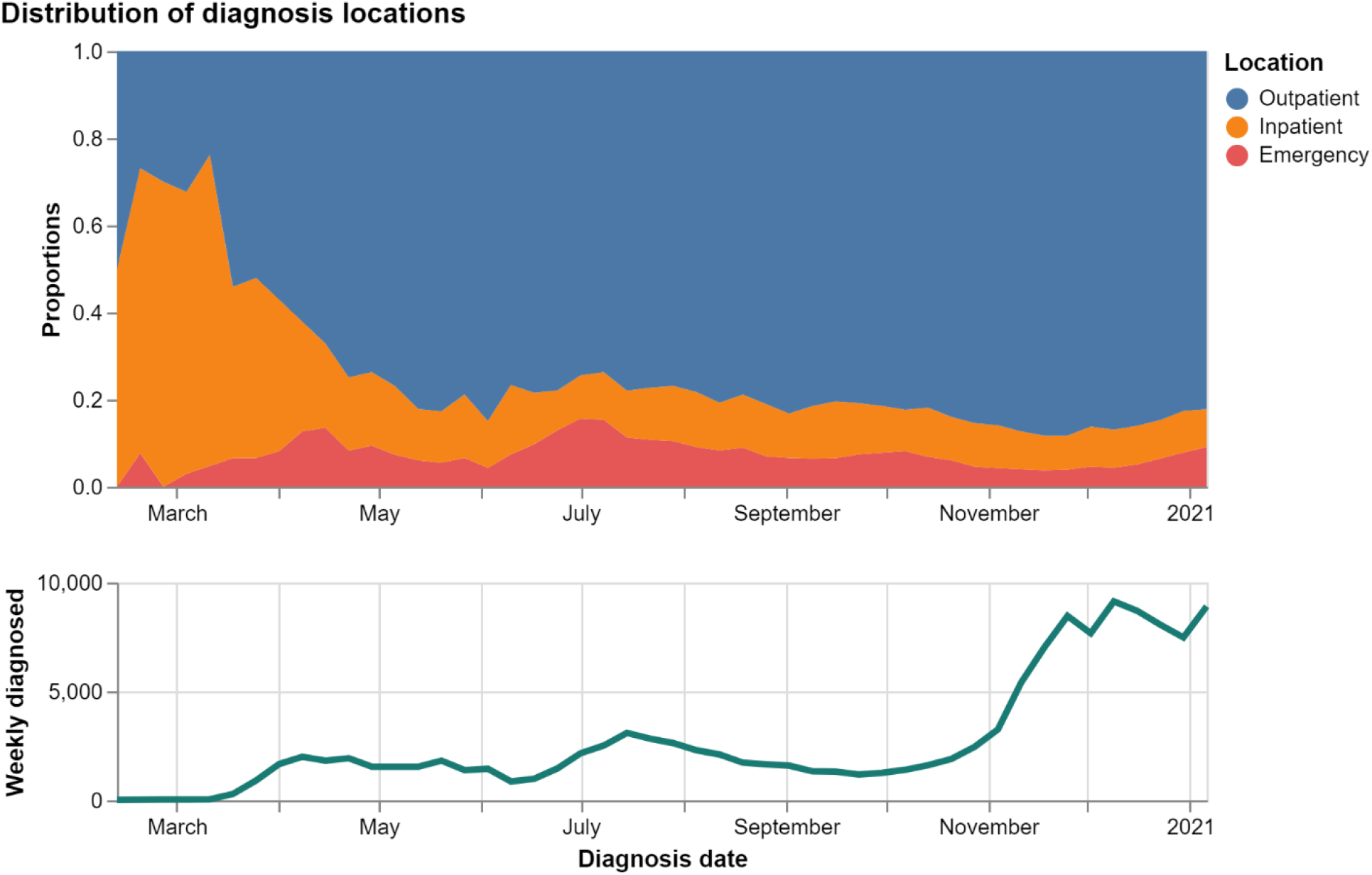
Temporal changes in SARS-CoV-2 diagnosis. Top panel shows the clinical setting in which participants’ first diagnosis was made. Over time as preparedness improved, more diagnoses are made in outpatient set tings (blue) than in inpatient (orange) or emergency room (red) setting. Bottom panel shows the number of new cases in every week – three surges are visible starting mid-March, mid-June, and October.

The top panel of Figure 2, presents the proportion of the medical settings in which individuals had their first record of infection. Early in the outbreak, a larger proportion of detections occurred in inpatient or emergency room setting. However, since late March more cases were detected in an outpatient setting, and this trend is largely continuing throughout the year. Overall, a larger proportion of diagnosis records originated from hospitals relative to outpatient settings in the first surge. This could be attributed to several processes: First, as preparedness increased testing have become more available to a broader population. Second, the virus was being introduced into a younger, possibly less susceptible, population, causing milder symptoms (if any) that did not require inpatient care. Third, as awareness increased in the population, individuals with suspected exposure were instructed not to arrive in the ER without being tested first ^13^.

### Temporal patterns of demographical and clinical characteristics

Figure 3 shows how the characteristics of individuals diagnosed throughout the pandemic change over time (grey dashed line). It further separates those detected with SARS-CoV-2 into two mutually exclusive groups – those who subsequently suffered a severe health event, and those who did not (red and blue lines, respectively). The characteristics presented are average age, proportion of sex, and average number of preexisting conditions, bound by a 95% confidence interval band.

**Figure 3.**
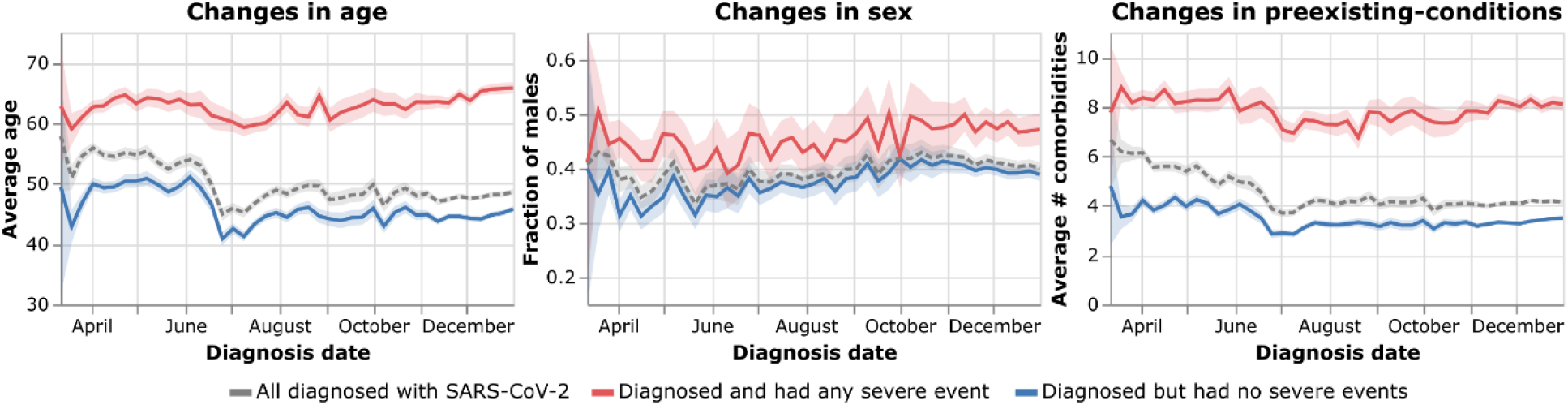
Temporal changes in background characteristics of individuals diagnosed with SARS-CoV-2. X-axis shows date of first SARS-CoV-2 diagnosis. Left panel: changes in average age. Middle panel: changes in proportion of males. Right panel: changes in average number of preexisting conditions. Grey dashed line presents all individuals diagnosed with SARS-CoV-2. Among them, infectees with co-occurring severe health events (red line) are constantly older and with more preexisting conditions than the complementary subgroup - those who had not suffered from severe events (Blue line). Error bands depict 95% confidence interval around the weakly mean.

Those suffering from severe health events are consistently older and with more preexisting conditions on average, consistent with both measures being positively correlated. Additionally, while the pooled averages of everyone detected show a decrease over time, the measures of the subgroup experiencing health events are more stable. The proportion of males detected with SARS-CoV-2 shows a gradual average increase over time, though not statistically significant. While the fraction of females among infected was higher than males, the fraction among those with co-occurring severe illness was lower and closer to the fraction of females in the general population (Table 1).

### Association between infection and severe events in hospitalized patients

Availability of SARS-CoV-2 tests and non-random testing policies may induce spurious correlations between SARS-CoV-2 detection and severe health events. Therefore, to avoid testing bias, we further focused on individuals who were admitted to emergency room or inpatient hospitalization starting from the second surge, assuming most patients in these settings were likely to be tested.

The dynamics of the incidence for each of the events exhibits a sharp increase at the day of SARS-CoV-2 detection (Supp-Figure 2). This may hint that patients were either detected before encountering the provider addressing the severe event, or that the severe event triggered an encounter which in turn led to an unrelated SARS-CoV-2 detection. Therefore, to avoid such cases, in the following association analysis, follow-up starts at the fourth day of admission and continues for 30 days after admission, to increase the likelihood that the events occurred *after* SARS-CoV-2 infection and within the clinically relevant time frame following detection.

Out of individuals with hospital encounters between June 7^th^, 2020 and November 18^th^, 2020, 15,486 patients had a positive SARS-CoV-2 test in the 14 days prior to hospitalization or within 3 days of hospitalization (denoted hospital-detected, Figure 1) and 410,502 did not have any known positive SARS-CoV-2 tests prior to hospitalization or until at least 14 days after follow-up, which is 30 days after the encounter (denoted hospital-control, Figure 1). A comparison of the prevalence of pre-existing conditions for individuals in these two sub-groups shows a high similarity between the groups (Table 2, Supp-Figure 1), especially compared to the base population (Table 1).

**Table 2.** Characteristics of participants in the hospital-encounter cohort, separated to those detected with SARS-CoV-2 and the rest.

|  | General Population |  | SARS-2 diagnosed |  |
| --- | --- | --- | --- | --- |
|  | n=410,502 | % | n=15,486 | % |
| **Demographics** |  |  |  |  |
| Age (Mean, STD) | 49.4 | 22.3 | 51.0 | 20.3 |
| **Sex** |  |  |  |  |
| Female | 251,478 | 61.3% | 8,803 | 56.8% |
| Male | 159,024 | 38.7% | 6,683 | 43.2% |
| **Race** |  |  |  |  |
| African American | 127,383 | 31.0% | 5,788 | 37.4% |
| Asian | 3,298 | 0.8% | 162 | 1.0% |
| Caucasian | 256,640 | 62.5% | 8,422 | 54.4% |
| Hispanic/Latino | 82 | 0.0% | 10 | 0.1% |
| Multi-racial | 7,758 | 1.9% | 440 | 2.8% |
| Other | 5,577 | 1.4% | 307 | 2.0% |
| Refused to classify | 3,645 | 0.9% | 194 | 1.3% |
| Missing | 6,119 | 1.5% | 163 | 1.1% |
| **Ethnicity** |  |  |  |  |
| Declined | 2,675 | 0.7% | 100 | 0.6% |
| Hispanic | 15,478 | 3.8% | 1,079 | 7.0% |
| Non-Hispanic | 285,292 | 69.5% | 9,842 | 63.6% |
| Other | 88,234 | 21.5% | 3,911 | 25.3% |
| Missing | 18,823 | 4.6% | 554 | 3.6% |
| **Smoking** |  |  |  |  |
| Never | 160,437 | 39.1% | 6,914 | 44.6% |
| Former | 76,573 | 18.7% | 3,042 | 19.6% |
| Current | 61,903 | 15.1% | 1,407 | 9.1% |
| Missing | 111,589 | 27.2% | 4,123 | 26.6% |
| **Insurance** |  |  |  |  |
| Medicaid | 54,261 | 13.2% | 1,793 | 11.6% |
| Medicare | 84,021 | 20.5% | 2,701 | 17.4% |
| Other | 4,476 | 1.1% | 259 | 1.7% |
| Other public | 4,409 | 1.1% | 167 | 1.1% |
| Private | 143,152 | 34.9% | 5,213 | 33.7% |
| Research | 0 | 0.0% | 0 | 0.0% |
| Self-pay | 70,738 | 17.2% | 2,835 | 18.3% |
| Missing | 120,183 | 29.3% | 5,353 | 34.6% |
| **Comorbidities** |  |  |  |  |
| Alcohol substance abuse | 43,716 | 10.6% | 1,115 | 7.2% |
| **Cardiac disorder** |  |  |  |  |
| Acute MI | 11,924 | 2.9% | 499 | 3.2% |
| Carditis cardiomyopathy | 17,524 | 4.3% | 755 | 4.9% |
| Cardiac arrest | 1,931 | 0.5% | 90 | 0.6% |
| CHF | 37,776 | 9.2% | 1,641 | 10.6% |
| Conduction | 26,161 | 6.4% | 1,018 | 6.6% |
| Coronary | 49,884 | 12.2% | 1,979 | 12.8% |
| Dysrhythmias | 95,717 | 23.3% | 3,957 | 25.6% |
| Pulmonary | 20,275 | 4.9% | 939 | 6.1% |
| Valve disorders | 31,041 | 7.6% | 1,037 | 6.7% |
| Cerebrovascular disease | 39,881 | 9.7% | 1,451 | 9.4% |
| Delirium dementia cognitive disorders | 15,066 | 3.7% | 810 | 5.2% |
| Diabetes | 113,991 | 27.8% | 5,212 | 33.7% |
| Epilepsy and convulsions | 17,940 | 4.4% | 554 | 3.6% |
| Gastrointestinal disorders | 91,361 | 22.3% | 3,178 | 20.5% |
| Hematologic disorders | 104,257 | 25.4% | 4,409 | 28.5% |
| Hematologic neoplasms | 9,059 | 2.2% | 329 | 2.1% |
| Hepatitis | 12,152 | 3.0% | 345 | 2.2% |
| HIV | 2,218 | 0.5% | 85 | 0.5% |
| Hypertension | 180,827 | 44.1% | 7,500 | 48.4% |
| Immunity disorders | 4,872 | 1.2% | 222 | 1.4% |
| Lipid metabolism disorders | 143,435 | 34.9% | 5,838 | 37.7% |
| Liver disorders | 43,248 | 10.5% | 1,748 | 11.3% |
| Malignant solid neoplasms | 70,739 | 17.2% | 2,099 | 13.6% |
| Migraine | 34,797 | 8.5% | 1,249 | 8.1% |
| Multiple sclerosis | 2,727 | 0.7% | 90 | 0.6% |
| Musculoskeletal connective tissue disorders | 25,305 | 6.2% | 843 | 5.4% |
| Neurological disorders | 69,805 | 17.0% | 2,643 | 17.1% |
| Nutritional deficiencies | 96,799 | 23.6% | 4,106 | 26.5% |
| Obesity | 119,147 | 29.0% | 5,467 | 35.3% |
| Pancreatic disorders | 10,541 | 2.6% | 274 | 1.8% |
| Parkinson's disease | 3,000 | 0.7% | 129 | 0.8% |
| Peripheral and visceral vascular disease | 36,387 | 8.9% | 1,334 | 8.6% |
| Psychiatric disorders | 139,213 | 33.9% | 4,841 | 31.3% |
| Pulmonary disorder |  |  |  |  |
| Asthma | 49,765 | 12.1% | 2,005 | 12.9% |
| COPD bronchiectasis | 20,941 | 5.1% | 811 | 5.2% |
| Lung disease due to external agents | 934 | 0.2% | 52 | 0.3% |
| Pleurisy pneumothorax | 16,012 | 3.9% | 677 | 4.4% |
| Pulmonary fibrosis | 4,189 | 1.0% | 163 | 1.1% |
| Respiratory failure | 27,477 | 6.7% | 2,690 | 17.4% |
| Renal disorders | 72,073 | 17.6% | 3,260 | 21.1% |
| Respiratory infections | 47,843 | 11.7% | 4,818 | 31.1% |
| Skin infections | 46,357 | 11.3% | 1,378 | 8.9% |
| Thyroid disorders | 63,279 | 15.4% | 2,325 | 15.0% |
| Outcomes |  |  |  |  |
| Cardiovascular | 12,670 | 3.1% | 637 | 4.1% |
| Hematologic immune | 13,459 | 3.3% | 790 | 5.1% |
| Hepato | 607 | 0.1% | 18 | 0.1% |
| Neurological | 8,999 | 2.2% | 466 | 3.0% |
| Renal | 17,888 | 4.4% | 1,330 | 8.6% |
| Respiratory bacterial pneumonia | 1,056 | 0.3% | 268 | 1.7% |
| Respiratory | 32,645 | 8.0% | 4,281 | 27.6% |
| Respiratory viral pneumonia | 119 | 0.0% | 2,960 | 19.1% |
| Sepsis | 6,489 | 1.6% | 550 | 3.6% |
| **Treatments** |  |  |  |  |
| **Anticoagulants** | 80,173 | 19.5% | 3,022 | 19.5% |
| **Convalescent plasma therapy** | 809 | 0.2% | 288 | 1.9% |
| **Cytokine inhibitors** | 221 | 0.1% | 119 | 0.8% |
| **Dexamethasone (and other corticosteroids)** | 115,623 | 28.2% | 5,434 | 35.1% |
| **Hydroxychloroquine** | 4,171 | 1.0% | 145 | 0.9% |
| **Lopinavir and ritonavir** | 13 | 0.0% | 2 | 0.0% |
| **Remdesivir** | 1,005 | 0.2% | 2,443 | 15.8% |
| **Tracheostomy** | 1,245 | 0.3% | 84 | 0.5% |
| **Ventilation moderate** | 22,844 | 5.6% | 1,199 | 7.7% |
| **Ventilation severe** | 3,620 | 0.9% | 322 | 2.1% |
| **Ventilators and other respiratory support devices** | 9,119 | 2.2% | 490 | 3.2% |
| **Outcomes baseline** |  |  |  |  |
| **Cardiovascular (30 days)** | 12,051 | 2.9% | 551 | 3.6% |
| **Hematologic immune (30 days)** | 11,937 | 2.9% | 692 | 4.5% |
| **Hepato (30 days)** | 515 | 0.1% | 20 | 0.1% |
| **Neurological (30 days)** | 8,623 | 2.1% | 367 | 2.4% |
| **Renal (30 days)** | 15,469 | 3.8% | 1,146 | 7.4% |
| **Respiratory bacterial pneumonia (30 days)** | 708 | 0.2% | 140 | 0.9% |
| **Respiratory (30 days)** | 44,775 | 10.9% | 4,292 | 27.7% |
| **Respiratory viral pneumonia (30 days)** | 118 | 0.0% | 1,728 | 11.2% |
| **Sepsis (30 days)** | 6,024 | 1.5% | 486 | 3.1% |

We tested for sensitivity to the lag between admission and follow-up start time (Supp-Figure 3). All outcomes were stable across these parameters, except cardiovascular which showed increasing odds ratios for greater lags. Associations where also stable for all outcomes replacing the 14 days lag from detection to admission by 7, 21, and 28 days but not on the extreme zero days lag. We also tested for sensitivity to the exclusion of 10,190 control patients by reweighting the regression with the inverse propensity to be censored, and results are stable (Supp-Figure 4).

An analysis of the association between SARS-CoV-2 detection and severe health events (Figure 4) shows significant positive associations between detection and most severe health events among hospitalized patients. The odds ratios are adjusted for baseline characteristics extracted for the pre-admission period and until the third day of admission (see Methods, unadjusted results are in Supp-Figure 5). We used viral pneumonia as a positive control, exhibiting an adjusted odds ratio of 594 (95% CI 492-718) (not shown in figures).

**Figure 4.**
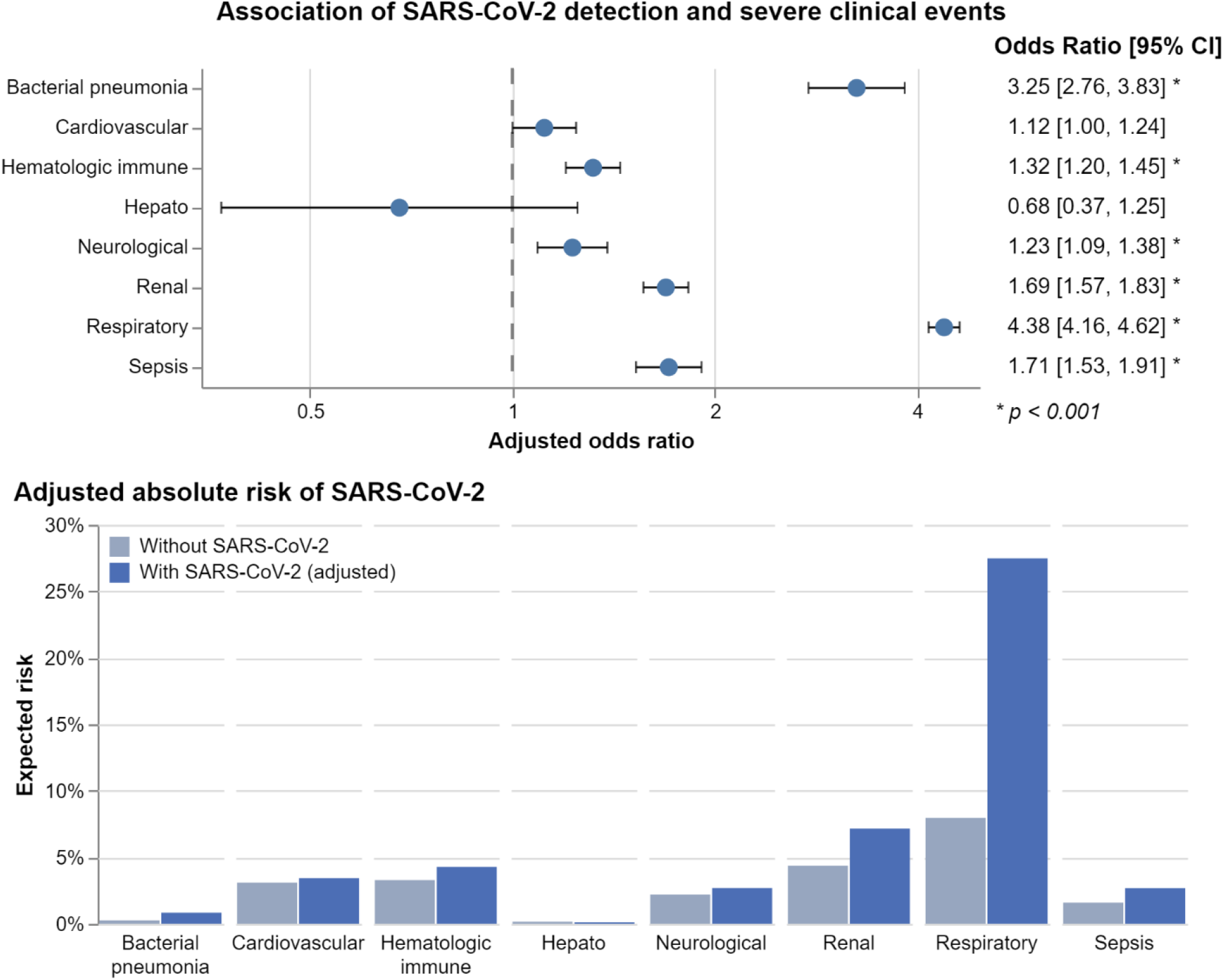
Association of SARS-CoV-2 detection with severe health events among hospitalized patients, adjusted for demographics, preexisting conditions and past events. Top panel presents the associations as odds ratios, with error bars representing 95% confidence intervals. Bottom panel converts odds ratios to absolute risks, showing baseline risk (grey) and the risk associated with SARS-CoV-2 infection in percentages. Overall, SARS-CoV-2 is associated with increased risk for most events.

Another, possibly more intuitive, way of presenting the results is using the absolute risk. For those hospitalized with no SARS-CoV-2 infection (hospital-control), this risk can be obtained directly from the data. The absolute risk for patients with SARS-CoV-2 (hospital-detected) is then calculated using the odds ratio that was obtained above (which is the ratio between the odds of the health event in the hospital-control and hospital-detected groups). Using this notation, for every 1000 patients in the hospital-control we observed 80 with severe respiratory events compared with 275 for every 1000 in the hospital-detected cohort. Similarly, these numbers are 16 vs. 27 cases of sepsis, 44 vs. 72 renal events, 33 vs. 43 hematologic events, 22 vs. 27 neurological events, and 3 vs. 8 bacterial pneumonia cases out of every 1000 patients in the hospital-control vs. the hospital-detected, respectively. The difference that was observed for cardiovascular events, of 31 vs. 34, was not statistically significant after multiple hypothesis correction (uncorrected P value of 0.05).

### Changes in treatment strategies over time

To further examine the reduction in the incidence of severe health events between surges (Supp-Figure 2), we examine the changes in treatments assignment for COVID-19 over time. Since not all treatments are available in all clinical settings (e.g., convalescent plasma), we further focus our cohort on inpatient hospitalizations only. Since a patient can get more than one treatment, Figure 5 presents the proportion of patients receiving a treatment among all the patients diagnosed within that week.

**Figure 5.**
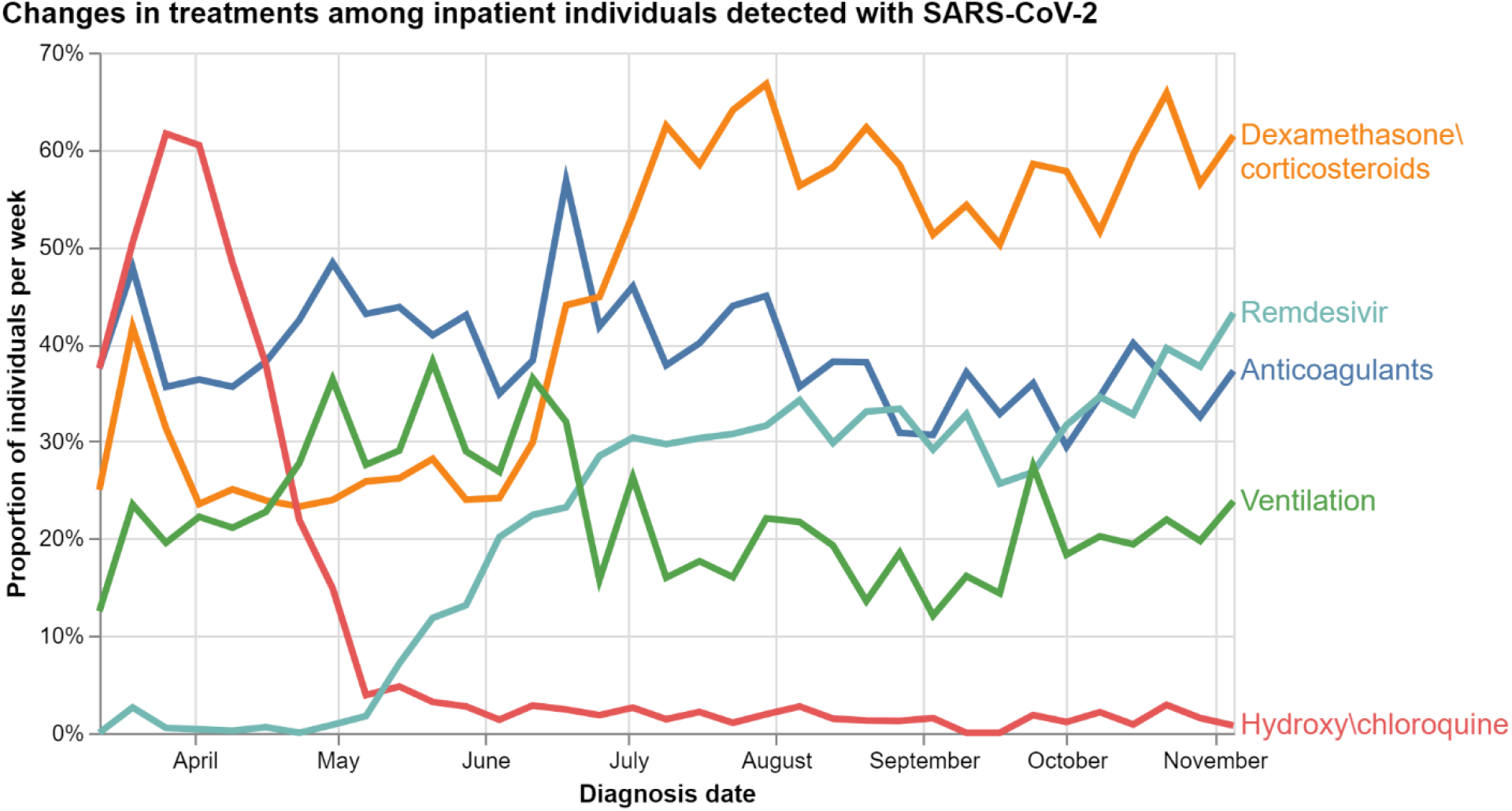
Temporal changes in treatments administered to hospital inpatient patients diagnosed with SARS-CoV-2. At each time point, Y-axis shows the percentage of people receiving a treatment among all patients diagnosed at that time point. For example, “62% of patients diagnosed with SARS-CoV-2 on the week of July 5^th^ were treated with Dexamethasone or other corticosteroids”. Each time-point sums to more than 100% because patients can receive multiple treatments. The plot shows increased use of dexamethasone and remdesivir over time, a sharp decline in use of chloroquine and hydroxychloroquine and a mild reduction in use of mechanical ventilation.

We see prominent use in chloroquine and hydroxychloroquine starting in March, co-occurring with the first publication showing potential benefits ^14^, followed by a sharp decrease co-occurring with the U.S. FDA cautioning against using the drug for COVID-19 in April ^15^ and later fully revoking its Emergency Use Authorization (EUA) in mid-June ^16^.

Not unexpectedly, we see a slow increase in the use of remdesivir starting in May after the U.S. FDA issues an EUA for its use in severe COVID-19 ^17^. Similarly, Dexamethasone use upticks in June, co-occurring with positive preliminary results from a randomized control trial ^18,19^. The trend for mechanical ventilation is less clear, and may reflect some lack of consensus around the practice ^20^.

## Discussion

This study examined the electronic health records of over 9 million individuals across the United States throughout 2020 to track the COVID-19 pandemic. We identified a set of eight severe events in multiple organs and focused on two questions. First, we examined 131,329 SARS-CoV-2 infected individuals to show the characteristics among those who also suffer severe health events were stable throughout the pandemic. Second, we used a set of 425,988 hospitalized patients to quantify the increase in incidence of severe respiratory, neurological, blood, kidney, heart, sepsis, and bacterial pneumonia events associated with SARS-CoV-2 diagnosis.

The weekly count of cases of SARS-CoV-2 manifests in three surges (sharp increase in cases) occurring on March, June, and October (Figure 2) all consistent with contemporaneous public reporting. Starting from the second surge and onwards, a younger population with fewer preexisting conditions, on average, is being diagnosed (Figure 3) – probably due to increased testing. However, when stratifiedon those also exhibiting severe health events, the subgroup is consistently older and with more preexisting conditions. Their characteristics also vary less over time, which might suggest that there was no observable change in pathogenicity of the virus during this study (e.g., due to selective pressures as it became more prevalent ^21^). However, the timeframe of the study limits us from observing an effect of the newer variants emerging in late 2020 ^22^.

The over-representation of infected females compared to males in the data does not indicate higher susceptibility to subsequent severe events. Rather, the proportion of diagnosed females suffering severe health events trends towards the rate observed in females in the entire data, suggesting that the risk for severe health events among males and females with SARS-CoV-2 infection is similar, and that there may have been some selection bias early in the pandemic ^23^. The sex disparity in our data may be attributed to over-utilization by females in specific healthcare services.

Despite that the COVID-19 outbreak impacted different regions across the U.S. at distinct times, data from various geo-diverse regions was aggregated for analysis and may introduce temporal variance. This aggregation also intermingles effects of non-pharmaceutical interventions taken in different places ^24,25^. However, the infection data extracted from the Explorys database follows similar trends to those observed in the entire US (Supp-Figure 7). This supports that the data, and its progression through time, is representative of the US population, and therefore the temporal changes we have outlined are generalizable to the US.

Incidence for all severe health events are higher in the first surge relative to the second surge (Supp- Figure 2). This could be contributed to any number of factors seen in the data: reduction in average age, reduction in average number of preexisting conditions (Figure 3), and changes in treatment strategies over time (Figure 5). Disentangling direct effects of background factors from treatments is an important subject for further research, and readers are advised not to draw causal conclusion from this apparent correspondence.

Our main analysis focused on 425,988 individuals in hospital encounters and has shown a positive association between SARS-CoV-2 detection and multiple non-pulmonary health events. As expected, the associations are smaller than the positive association of SARS-CoV-2 detection with pulmonary health events but are still statistically and practically significant. These results complement pathophysiological and small-scale studies as reviewed in Gupta et al. ^12^ with large-scale data-driven quantifications.

We focused on hospitalized patients from the second surge onwards. While this can’t account for admission hesitancy during a pandemic, it can help reduce two sources of bias we can’t control for in our data: non-pharmaceutical intervention policies in different geographic locations that affect infection susceptibility; and testing strategies that can favor at-risk individuals, since admitted patients are almost certain to be tested. Consequently, the baseline characteristics among those with and without diagnosis are quite similar between the two groups in the hospitalization cohort (Table 2), much more sothan in the base population (Table 1). Although it may introduce selection bias ^26^, this is likely to reduce confounding when measuring the statistical contribution of infection to severe events. Focusing on hospitalizations also mitigates gaps in the data because it increases the likelihood of a more complete longitudinal data set, since the Explorys database is focused on aggregating across specific health care systems rather than specific patients. Our focus on the second surge and onward allows covering a broader population as testing expanded to include a younger population with milder or no symptoms (Figure 3) which represents the overall population better. Additionally, as clinical care improved with time (higher incidence of severe health events during the first surge, see Supp-Figure 2), focusing on hospitalization after the second surge reduces the chance of positive associations being due to suboptimal clinical practices. Lastly, the timeframe of the study is prior to mass distribution of vaccines and therefore unaffected by them.

Since the outcomes are measured from electronic health records, they are less subjective than previous questionnaire-based studies on non-respiratory illness ^27^. On the other hand, the dataset is not fully linked to mortality records. This may cause underestimation of the risk due to censoring if, for example, hospitalized patients with SARS-CoV-2 have a higher propensity of death compared to others. However, we defined the severe health events to be severe enough so that it should be reasonable to assume that most hospital deaths are preceded by one of these events.

Focusing on hospitalizations and starting follow-up three days after admission improves our confidence that the severe events did not occur prior to infection. Positive associations between SARS-CoV-2 detection and severe events (Figure 4) can have several explanations. First, background comorbidities are considered risk-factors for COVID-19. Second, non-COVID medical conditions can trigger hospitalization, which in turn triggers testing for SARS-CoV-2. Third, direct viral pathogenic effects or indirect effects of the immune response may promote adverse events in different systems. Forth, there may be common factors that affect both susceptibility to COVID-19 as well as to other conditions. To disentangle some of the factors and isolate the contribution of infection, we adjusted for many risk factors and focused the analysis on detections that necessitate hospital admissions and that also occur sufficient time after hospital admission. However, as residual biases may be still prevalent, additional data source are needed to better clarify the cause of the inferred associations.

## Conclusions

While we could not determine an association between severe liver events and SARS-CoV-2 infection (95% CI of adjusted-OR [0.37, 1.25]), and cardiovascular illness has borderline statistically significant (adjusted-OR 1.12, *p*=0.05 before adjusting to multiple hypotheses), we saw SARS-COV-2 significantly associated with increase in risk of severe blood, kidney, neurological, and respiratory events among hospitalized patients. While the absolute risk of non-respiratory events may not be large from an individual perspective (0.5 to 3 percentage point change), the severity of these events renders these results non-negligible. Regardless, it is significant from a healthcare perspective as it represents thousands of potential patients across the US. For severe respiratory events, the risk increase associated with SARS-CoV-2 is very high even in absolute terms (8% to 28%). Additionally, all outcomes that presented high increase in relative risk (22% to 69% more likely) should be of importance for the medical community for public health considerations.

This study used millions of electronic health records to characterize the temporal changes among individuals infected with SARS-CoV-2, compare them to a baseline population and quantify the association between infection and both respiratory and non-respiratory severe health events. We found the risk increase associated with SARS-CoV-2 infection to be significant both statistically and practically. The risk consistently higher for older population with preexisting health conditions throughout the pandemic.

We believe this study sheds more light on how we understand the pandemic quantitively and help the medical community grasp the risk of various manifestation of SARS-CoV-2.

## Methods

### Cohort development

We constructed a *base population cohort* of individuals that had an encounter during 2019, and who also had at least one year of history on January 1, 2020. Events were extracted up to January 6, 2021.

The *hospital encounter* cohort consists of individuals who were admitted to hospital or ER for any reason between June 7 and November 18. We excluded individuals who had an infection more than 14 days prior to hospitalization. We define the follow-up period as the 30 days starting at 4-days after hospitalization. The main analysis also excluded patients who had SARS-CoV-2 detected within the follow up time plus an additional 14 days to account for delayed diagnosis. See Figure 1 for a graphical representation of the inclusion/exclusion criteria.

### SARS-CoV-2 infection

SARS-CoV-2 detection identified by both diagnostic and lab-test results codes. Specifically: (1) the official code ICD-10 code U07.1; (2) LOINC codes of SARS-Cov-2 tests with positive results; (3) Textual diagnosis descriptions such as “2019 novel coronavirus detected that were manually reviewed for accuracy; and (4) ICD, LOINC or text codes of other coronaviruses intersected with respiratory problems (see Data supplement A).

### Outcomes

Following Gupta et al. ^12^, we extracted indictors for respiratory events as well as non-pulmonary manifestations including cardiovascular, hematologic/immune, renal, neurological, hepatic and sepsis. We identified severe events of these manifestations by medical expertise review and mapped them into diagnostic, lab, and procedure codes (Table 3).

**Table 3.** Definitions of severe clinical events.

| Name | Events |
| --- | --- |
| **Respiratory** | **According to NIH guidelines:**<sup>28</sup> respiratory frequency >30 breaths per minute, SpO2 <94% on room air at sea level, ratio of arterial partial pressure of oxygen to fraction of inspired oxygen (PaO2/FiO2) <300 mmHg, or lung infiltrates >50% (we use viral pneumonia as a proxy for lung infiltrates detection).<br>Additionally, we use the following indicators:<br>ARDS, respiratory arrest, respiratory failure, ventilation: non-invasive/invasive, tracheostomy, ECMO, respiratory acidosis, acidosis of unspecified type. |
| **Bacterial pneumonia** | Bacterial pneumonia |
| **Hematologic/immune** | Arterial thrombotic complications: myocardial ischemia, ischemic stroke, acute limb, and mesenteric ischemia; Pulmonary embolism; Hypotension; Shock |
| **Sepsis** | Sepsis, septicemia, septic shock |
| **Cardiovascular** | Myocardial ischemia, myocardial infarction, myocarditis; Heart failure, cardiac arrest; Arrhythmia: torsades de pointes and pulseless electrical activity; cardiogenic shock; Pulmonary embolism; Need for CPR |
| **Renal** | Acute kidney injury requiring dialysis; Metabolic acidosis pH under 7.1, acidosis of unspecified type; Clotting of extracorporeal circuits used for renal replacement therapy |
| **Hepatobiliary** | Severe acute liver injury |
| **Neurological** | Encephalopathy<sup>1</sup>; Stroke; Ischaemic strokes; Neurogenic shock<sup>29</sup> |

<sup>1</sup> Meningoencephalitis, hemorrhagic posterior reversible encephalopathy syndrome, acute necrotizing encephalopathy, including the brainstem and basal ganglia, Guillain-Barré syndrome.

### Treatments

Treatment examined were anticoagulants usage, dexamethasone or other corticosteroids, hydroxychloroquine or chloroquine, remdesivir, cytokine inhibitors, lopinavir or ritonavir, convalescent plasma as well as ventilation and tracheostomy. These treatments were encoded either in ICD-9-CM, ICD-10-PCS, SNOMED, CPT procedure codes or RXCUI for drugs (see Data supplement B).

### Preexisting conditions

Pre-existing conditions were extracted based on *clinical classifications software* (CCS) codes ^30^. These included known risk-factors for COVID-19 such as hypertension, diabetes type 2, heart conditions as well as other conditions, summing up to 44 comorbidities. Some CCS codes were grouped into a single code (e.g., different cancers were all treated as a single neoplasm category). CCS codes were then mapped into corresponding ICD-9 CM and SNOMED CT codes. A detailed mapping can be found in (see Data supplement C).

A person was considered diagnosed with a comorbidity if they either had diagnostic code anytime in 2019, or if they had a record of an unresolved problem (either self-reported or filled in by a provider) in their problem list with a corresponding comorbidity code as above.

### Demographic variables

Demographic variables used are age (in years), sex (binary), insurance type (7 categories), and self-reported race (7 categories), ethnicity (4 categories), and smoking status (3 categories). To reduce missing data, and since demographic characteristics tend to vary less, they were extracted from records of up to 10 years before the 2020 index date.

### Temporal analysis

Number of people were aggregated by week to avoid the observed dip in counts during weekends. Python 3.6 with Pandas 0.23.4 ^31^ were used for data manipulation and plotting was done with Altair 4.1.0 ^32^.

### Association analysis

A preliminary exploration with Kaplan-Meyer plots of severe-events showed a large proportion of events. Therefore, to reduce the likelihood of considering encounter reasons as outcomes, we started the follow-up period 4 days after admission.

We applied multi-variable logistic regression for every severe event of interest in a follow-up period that begins at the fourth day after admission and last for 30 days. The exposure variable is an indicator of SARS-CoV-2 detection in the period that spans 14 days before and up to the third day after admission day. The three-day grace period was taken to account for diagnosis latency. Control variables included demographic and clinical features, and indicators of severe events in the preceding 30 days.

We performed sensitivity analysis for both design decisions: ranging the follow-up start time from zero to four days and ranging detection lag from zero to four weeks. Finally, we tested sensitivity to loss to follow-up using inverse probability of censoring weighting, fitting a logistic regression, using the same predictors, to predict late detection among the control patients.

## Data Availability

Data supporting the findings of this study are available from IBM Explorys but were used under license for the current study and restrictions may apply to their availability.

## Acknowledgment

The authors thank IBM Research for facilitating this research. Especially to Belinda Ann Perez and Tammy Allen from IBM Watson Health for their assistance in understanding the clinical codes; Ben Kolt from IBM Explorys for his help understanding the Explorys Database; To the Explorys IT team for their timely problem fixing; and to the Machine Learning for Healthcare team at IBM Research Haifa for their discussions and insights.

## Contributors

**Conceptualization:** TEH, YS, EK, MN, AJ, MRZ. **Data curation**: MN, SR, MC, AP, **Formal Analysis**: TEH, EK. **Methodology**: TEH, EK, YS, MN. **Software**: TEH, EK, MN, SR. **Supervision**: AP, AJ, TP, MRZ, YS. **Visualization**: EK. **Writing – original draft**: TEH, EK, YS. **Writing – review & editing**: TEH, EK, YS, AP, AJ, TP, MN, SR, MC, MRZ.

## Research in context

### Evidence before this study

SARS-CoV-2 is a respiratory virus. Nevertheless, since early in the pandemic, there have been reports of COVID-19 patients suffering from non-respiratory complications. SARS-CoV-2 has been found to affect multiple organ systems, such as the cardiovascular, hematologic, immune, hepatic, renal and neurological systems. These correlations where even hypothesized to be driven by underlying risk factors confounding the associations. Studies have hypothesized about the physiological mechanisms and biological pathways from which the virus causes extrapulmonary manifestations. However, to the best of our knowledge, a systematic, big-data-based quantification of the risk for severe events in such systems associated with the virus, has not-yet been established.

### Added value of this study

We used 9,344,021 electronic health records from the IBM Explorys database to construct a cohort of hospitalized patients and a general cohort of individuals detected with SARS-CoV-2. We defined a set of severe health events in respiratory and non-respiratory organ systems using medical codes. We modelled the association between SARS-CoV-2 diagnosis and these severe outcomes to quantify the risk of infection. Associations, however, can rise spuriously since SARS-CoV-2 infection and severe events may be bi-directional and confounded by various factors. Therefore, we designed our inclusion/exclusion criteria to reduce selection bias and adjusted for multiple demographic factors and rich clinical history to reduce confounding bias between the infection and the outcomes. We found that among hospitalized patients with SARS-CoV-2 are expected to show 22% to 69% more severe extrapulmonary events than uninfected patients. We further examined how the risk changes over time for the general population to discover that people suffering from severe outcomes showed no observable change in demographic or clinical factors throughout most of the pandemic.

### Implications of all the available evidence

The ability to quantify the risk is an imperative step towards optimal risk management. Physicians now have both the physiological mechanism and a quantitative estimation of the risk of extrapulmonary manifestation associated with SARS-CoV-2 infection. This can further allow hospitals to better adjust for expected cases and let policy maker better direct resources based on quantitative assessments.

## Supplementary materials

**Supp-Figure 1.**
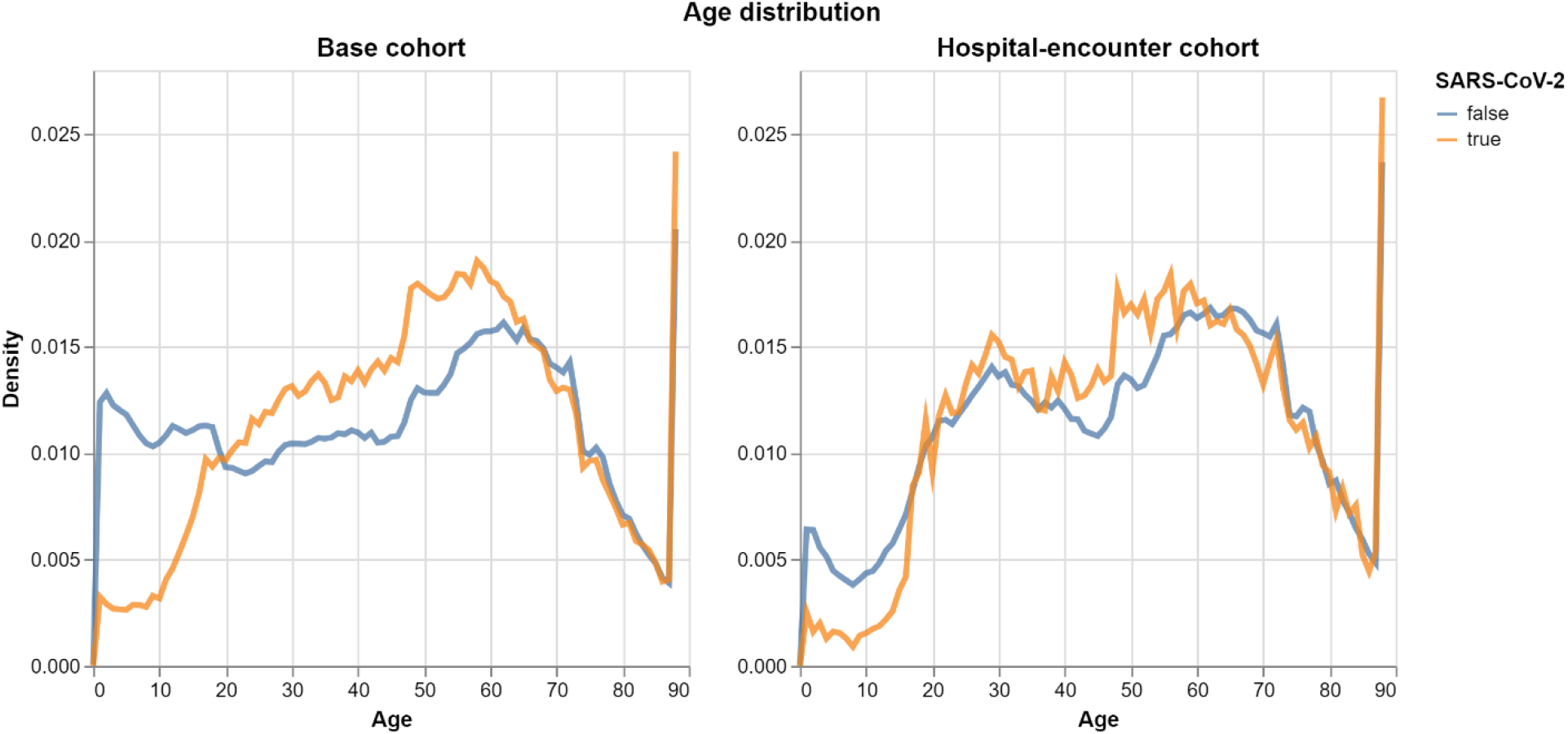
Age distribution of the participants in the study, in both base population cohort (left) and the hospital-encouter cohort (right), and separated to those detected with SARS-CoV-2 (orange) and the rest (blue). Ages above 90 are clipped.

**Supp-Figure 2.**
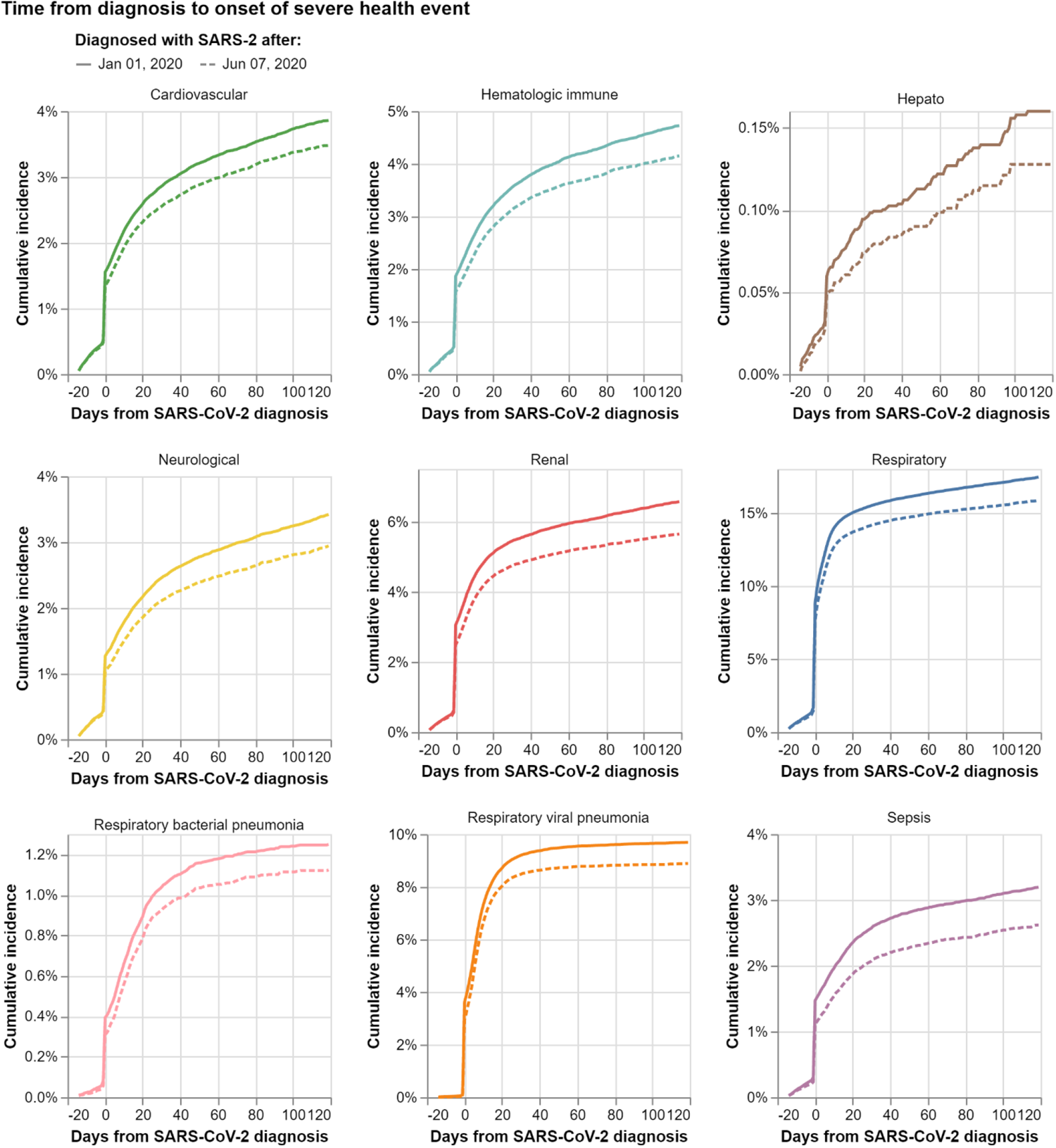
Cumulative incidence of severe health events in the base population detected with SARS-CoV-2. Estimated with Kaplan Meier. First surge (solid line) consistently presents higher incidence than second surge (dashed line). Index date is set at day of first SARS-CoV-2 diagnosis, the sharp rise suggests either patients were detected before an encounter with the provider that address the severe event, or that the severe event triggered an encounter which in turn led to a SARS -CoV-2 test; leading to the criteria in association analysis where events should occur after detection.

**Supp-Figure 3.**
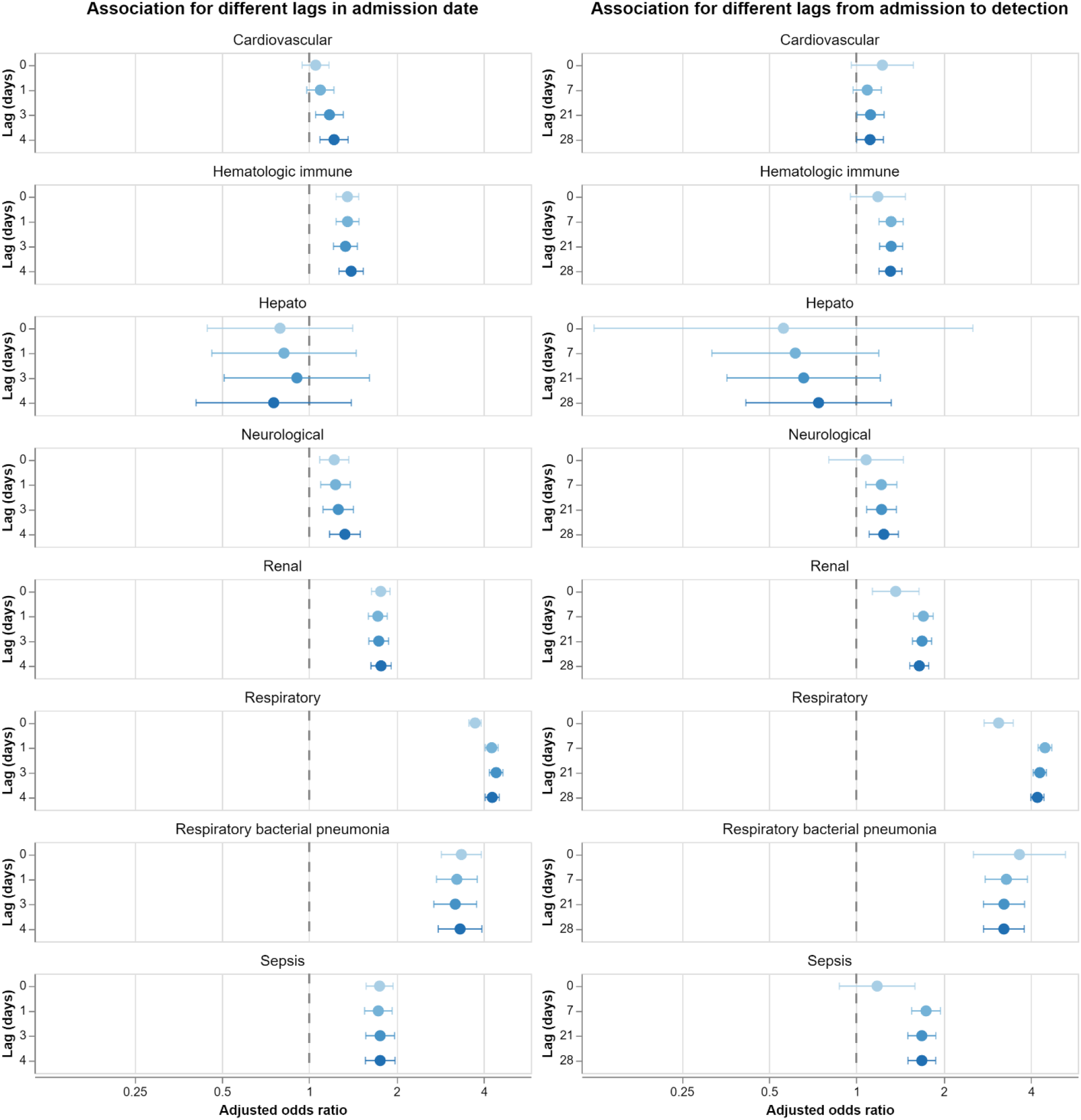
Sensitivity analysis of associations for different time-lag choices. Association between SARS-CoV-2 detection and severe health events, obtained from a regression model for various design choices. Left: Changes in the days between hospital admission date to the first day of follow-up. The larger the lag the greater the confidence that the patient was not admitted because of the event, but rather developed it after admission. Right: Changes in the maximum number of days from SARS -CoV-2 detection to admission in the hospital-detected group. Zero interval makes it harder to distinguish COVID from non-COVID causes for admission, on the other extreme, larger lags increase the likelihood for other non-COVID events to cause admission.

**Supp-Figure 4.**
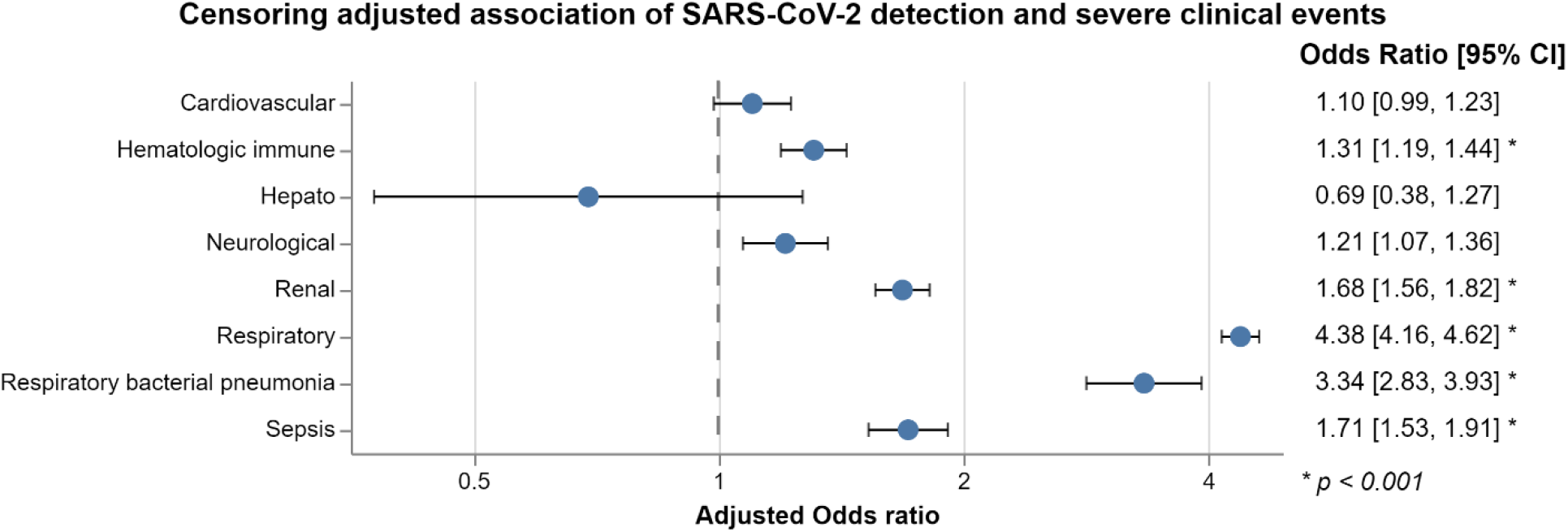
Risk-adjusted and censoring-adjusted associations between SARS-CoV-2 detection and severe health events. Associations adjusted for demographics, medical history, and baseline events through logistic regression, but also weighted by the inverse probability of censoring – control samples that are diagnosed with SARS-CoV-2 long after admission and were excluded in the main analysis. Changes in associations between this and main analysis are de-minimis.

**Supp-Figure 5.**
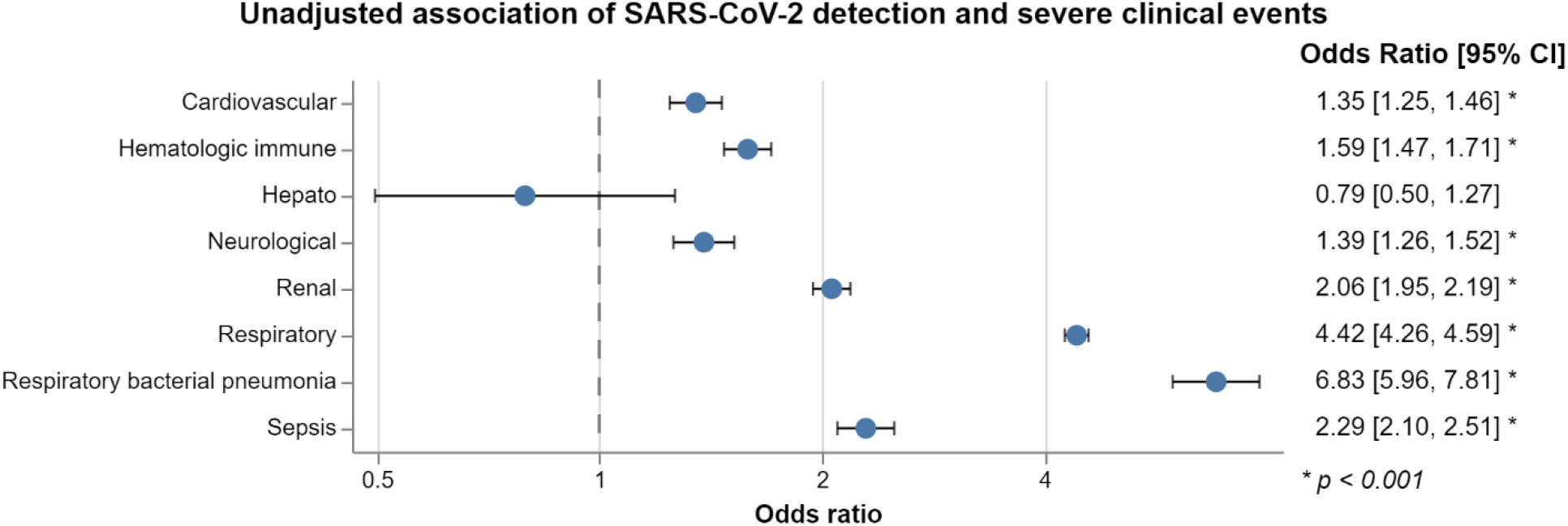
Unadjusted associations between SARS-CoV-2 detection and severe health events. Associations are not adjusted for demographics, medical history, and baseline events.

**Supp-Figure 6.**
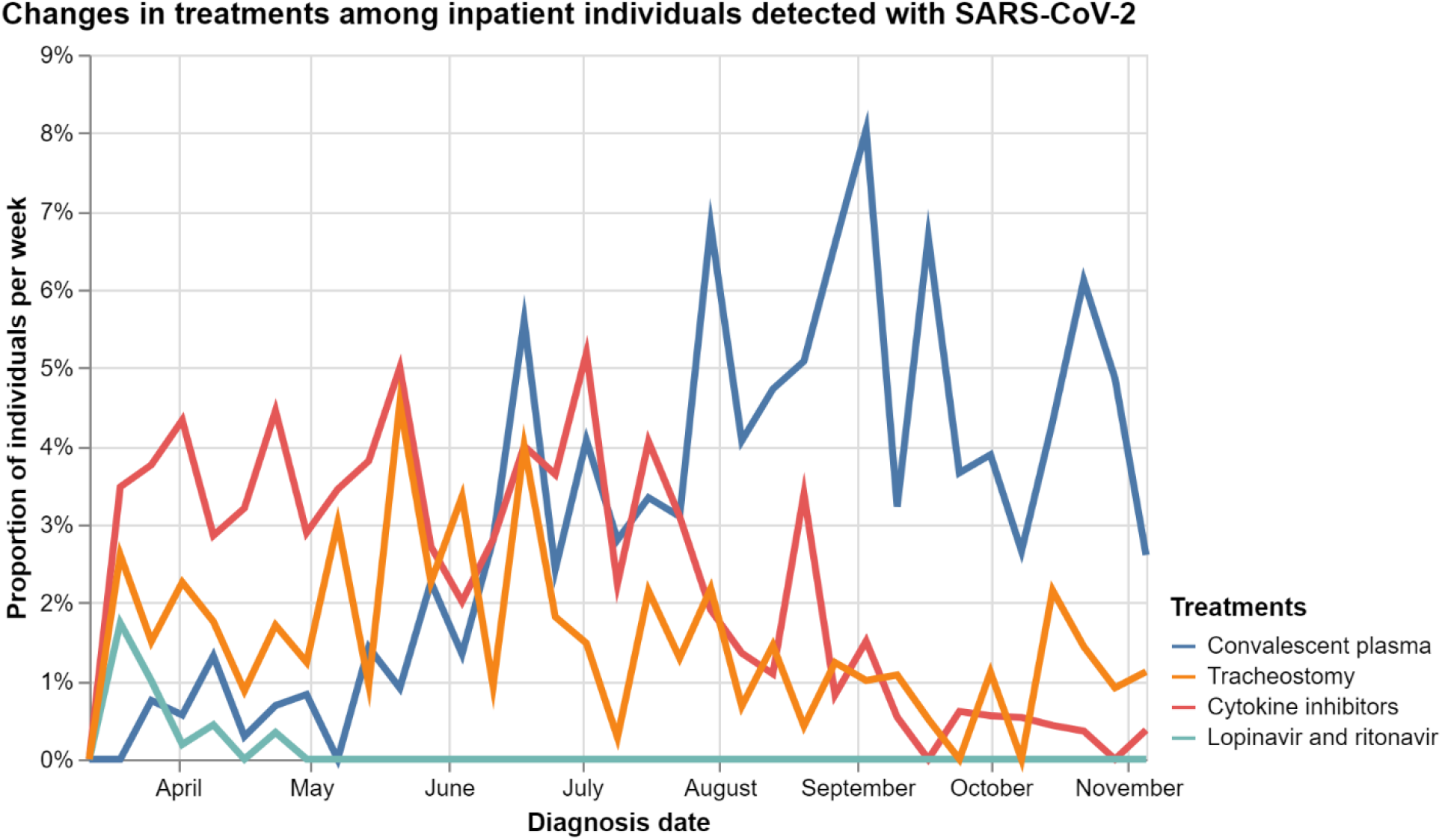
Temporal changes in treatments administered to hospital inpatient patients diagnosed with SARS-CoV-2. Same as Figure 4, but for less prevalent treatments.

**Supp-Figure 7.**
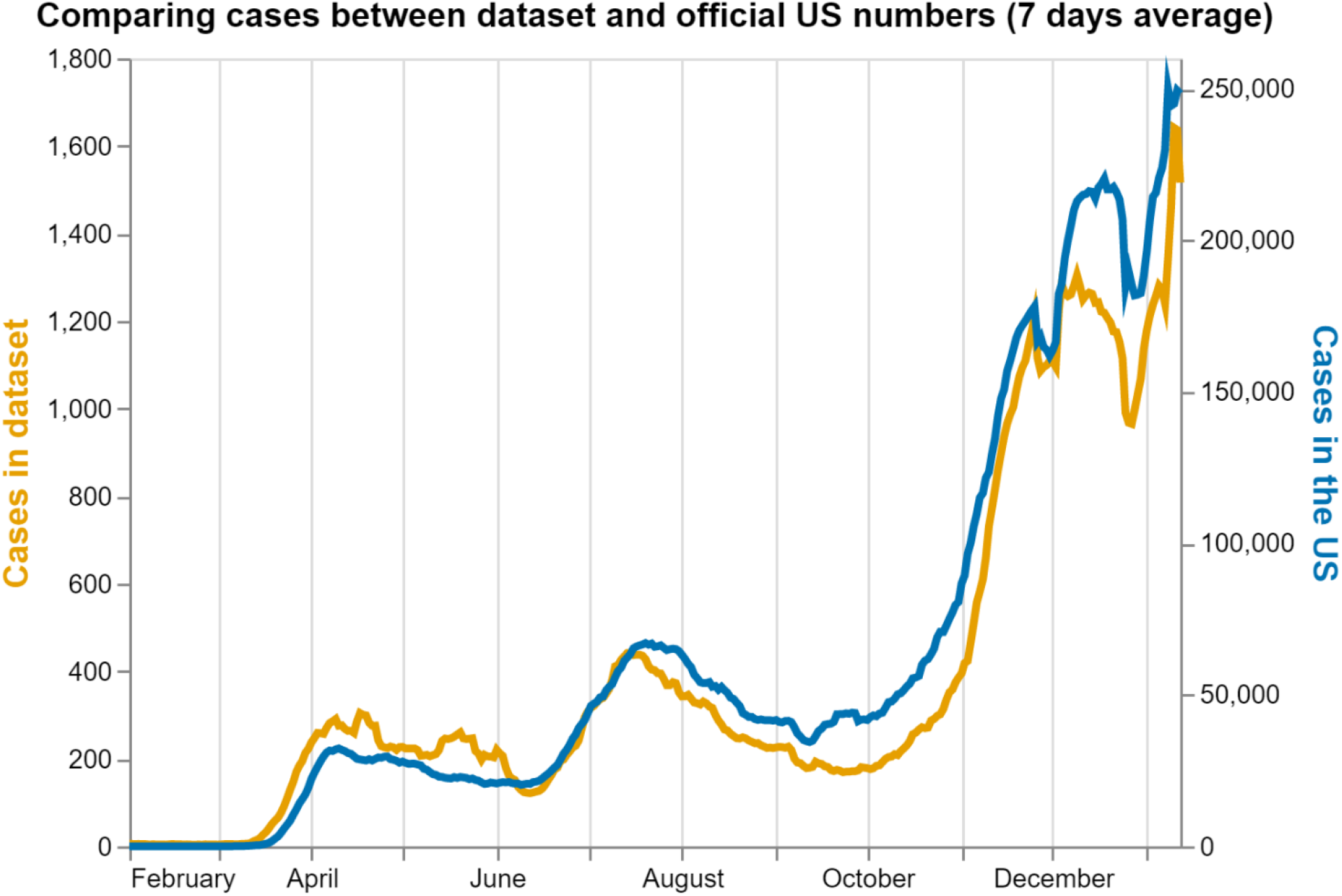
Trend of new cases in the dataset matches that of the US. Number of newly diagnosed cases in the dataset (dark yellow) juxtaposed against official case numbers obtained from John Hopkins’ dashboard data (Dong, Du, and Gardner, 2020)

## References

1. World Health Organization. Pneumonia of Unknown Cause – China. 2020 Jan 5 https://www.who.int/csr/don/05-january-2020-pneumonia-of-unkown-cause-china/en/ [Accessed: 2020-12-15].

2. Holshue ML, DeBolt C, Lindquist S, Lofy KH, Wiesman J, Bruce H, et al. First case of 2019 novel coronavirus in the United States. New England Journal of Medicine. 2020.

3. Center for Disease Control. COVIDView Week 49. 2020 Dec 11 https://www.cdc.gov/coronavirus/2019-ncov/covid-data/pdf/covidview-12-11-2020.pdf [Accessed: 2020-12-15].

4. Richardson S, Hirsch JS, Narasimhan M, Crawford JM, McGinn T, Davidson KW, et al. Presenting characteristics, comorbidities, and outcomes among 5700 patients hospitalized with COVID-19 in the New York City area. JAMA. 2020; 323: 2052–2059.

5. Vahidy FS, Drews AL, Masud FN, Schwartz RL, Boom ML, Phillips RA, et al. Characteristics and outcomes of COVID-19 patients during initial peak and resurgence in the Houston metropolitan area. JAMA. 2020; 324: 998–1000.

6. Nie Y, Li J, Huang X, Guo W, Zhang X, Ma Y, et al. Epidemiological and clinical characteristics of 671 COVID-19 patients in Henan Province, China. International journal of epidemiology. 2020; 49: 1085– 1095.

7. Mizrahi B, Shilo S, Rossman H, Kalkstein N, Marcus K, Barer Y, et al. Longitudinal symptom dynamics of COVID-19 infection. Nature communications. 2020; 11: 1–10.

8. Ayoubkhani D, Khunti K, Nafilyan V, Maddox T, Humberstone B, Diamond I, et al. Epidemiology of post-COVID syndrome following hospitalisation with coronavirus: a retrospective cohort study. medRxiv. 2021.

9. Yanover C, Mizrahi B, Kalkstein N, Marcus K, Akiva P, Barer Y, et al. What Factors Increase the Risk of Complications in SARS-CoV-2–Infected Patients? A Cohort Study in a Nationwide Israeli Health Organization. JMIR public health and surveillance. 2020; 6: e20872.

10. Gross CP, Essien UR, Pasha S, Gross JR, Wang Sy, Nunez-Smith M. Racial and Ethnic Disparities in Population-Level Covid-19 Mortality. Journal of General Internal Medicine. 2020 Aug; 35: 3097– 3099.

11. Dyer O. Covid-19: Minorities account for 78% of US deaths in under 21s, says CDC. BMJ. 2020 Sep;: m3681.

12. Gupta A, Madhavan MV, Sehgal K, Nair N, Mahajan S, Sehrawat TS, et al. Extrapulmonary manifestations of COVID-19. Nature Medicine. 2020 Jul; 26: 1017–1032.

13. Cleveland Clinic. Covid-19 Testing. 2020 https://my.clevelandclinic.org/landing/medical-professionals-preparing-for-coronavirus#testing-tab[Accessed: 2020-12-15].

14. Gautret P, Lagier JC, Parola P, Hoang VT, Meddeb L, Mailhe M, et al. Hydroxychloroquine and azithromycin as a treatment of COVID-19: results of an open-label non-randomized clinical trial. International Journal of Antimicrobial Agents. 2020 Jul; 56: 105949.

15. The United States Food and Drug Administration. FDA cautions against use of hydroxychloroquine or chloroquine for COVID-19 outside of the hospital setting or a clinical trial due to risk of heart rhythm problems. 2020 Apr 24 https://www.fda.gov/drugs/drug-safety-and-availability/fda-cautions-against-use-hydroxychloroquine-or-chloroquine-covid-19-outside-hospital-setting-or [Accessed: 2020-12-15].

16. The United States Food and Drug Administration. Coronavirus (COVID-19) Update: FDA Revokes Emergency Use Authorization for Chloroquine and Hydroxychloroquine. 2020 Jun 15 https://www.fda.gov/news-events/press-announcements/coronavirus-covid-19-update-fda-wrevokes-emergency-use-authorization-chloroquine-and[Accessed: 2020-12-15].

17. The United States Food and Drug Administration. Coronavirus (COVID-19) Update: FDA Issues Emergency Use Authorization for Potential COVID-19 Treatment. 2020 May 1https://www.fda.gov/news-events/press-announcements/coronavirus-covid-19-update-fda-issues-emergency-use-authorization-potential-covid-19-treatment [Accessed: 2021-02-01].

18. The RECOVERY Collaborative Group. Dexamethasone in Hospitalized Patients with Covid-19 — Preliminary Report. New England Journal of Medicine. 2020 Jul.

19. Oxford University News Release. Low-cost dexamethasone reduces death by up to one third in hospitalised patients with severe respiratory complications of COVID-19. 2020 Jun 16 https://www.recoverytrial.net/files/recovery_dexamethasone_statement_160620_v2final.pdf [Accessed: 2020-12-15].

20. Torjesen I. Covid-19: When to start invasive ventilation is “the million dollar question”. BMJ. 2021 Jan;: 121.

21. van Dorp L, Acman M, Richard D, Shaw LP, Ford CE, Ormond L, et al. Emergence of genomic diversity and recurrent mutations in SARS-CoV-2. Infection, Genetics and Evolution. 2020 Sep; 83: 104351.

22. Leung K, Shum MHH, Leung GM, Lam TTY, Wu JT. Early transmissibility assessment of the N501Y mutant strains of SARS-CoV-2 in the United Kingdom, October to November 2020. Eurosurveillance. 2021 Jan; 26.

23. Cai H. Sex difference and smoking predisposition in patients with COVID-19. The Lancet Respiratory Medicine. 2020 Apr; 8: e20.

24. Suryanarayanan P, Tsou CH, Poddar A, Mahajan D, Dandala B, Madan P, et al. WNTRAC: AI Assisted Tracking of Non-pharmaceutical Interventions Implemented Worldwide for COVID-19. 2020 Sep 2.

25. Brauner JM, Mindermann S, Sharma M, Johnston D, Salvatier J, Gavenčiak T, et al. Inferring the effectiveness of government interventions against COVID-19. Science. 2020 Dec;: eabd9338.

26. Griffith GJ, Morris TT, Tudball MJ, Herbert A, Mancano G, Pike L, et al. Collider bias undermines our understanding of COVID-19 disease risk and severity. Nature Communications. 2020 Nov; 11.

27. Huang C, Huang L, Wang Y, Li X, Ren L, Gu X, et al. 6-month consequences of COVID-19 in patients discharged from hospital: a cohort study. The Lancet. 2021 Jan; 397: 220–232.

28. National Institutes of Health. Clinical Spectrum of SARS-CoV-2 Infection. 2020 Dec https://www.covid19treatmentguidelines.nih.gov/overview/clinical-spectrum/ [Accessed:2020-12-15].

29. Paterson RW, Brown RL, Benjamin L, Nortley R, Wiethoff S, Bharucha T, et al. The emerging spectrum of COVID-19 neurology: clinical, radiological and laboratory findings. Brain. 2020 Jul; 143: 3104–3120.

30. Healthcare Cost and Utilization Project (HCUP). HCUP CCS. 2017 Mar https://www.hcup-us.ahrq.gov/toolssoftware/ccs/ccs.jsp.

31. McKinney W. Data Structures for Statistical Computing in Python. In van der Walt S, Millman J, editors. Proceedings of the 9th Python in Science Conference; 2010. p. 56–61.

32. VanderPlas J, Granger B, Heer J, Moritz D, Wongsuphasawat K, Satyanarayan A, et al. Altair: Interactive Statistical Visualizations for Python. Journal of Open Source Software. 2018; 3: 1057.

33. Greenbaum NR, Jernite Y, Halpern Y, Calder S, Nathanson LA, Sontag DA, et al. Improving documentation of presenting problems in the emergency department using a domain-specific ontology and machine learning-driven user interfaces. International Journal of Medical Informatics. 2019 Dec; 132: 103981.

34. Gardner RL, Cooper E, Haskell J, Harris DA, Poplau S, Kroth PJ, et al. Physician stress and burnout: the impact of health information technology. Journal of the American Medical Informatics Association. 2018 Dec; 26: 106–114.

